# Effect of photobiomodulation to control pain after placement of elastomeric spacers: a randomized controlled study protocol

**DOI:** 10.1101/2023.10.02.23296466

**Authors:** Luis Eduardo Pascuali Moya, Rolf Wilhem Consolandich Cirisola, María Victoria García Olazabal, Laura Hermida Bruno, Federico Todeschini Safi, Lucia Piriz Trindade, Priscila Larcher Longo, Maria Cristina Chavantes, Ricardo Scarparo Navarro, Cinthya Cosme Gutierrez Duran, Kristianne Porta Santos Fernandes, Raquel Agnelli Mesquita Ferrari, Sandra Kalil Bussadori, Lara Jansiski Motta, Anna Carolina Ratto Tempestini Horliana

**Author notes:** **<u>Corresponding Author:</u>** (ACRTH). Universidade Nove de Julho (UNINOVE), São Paulo, Brazil (Luis Eduardo Pascuali Moya, Rolf Wilhem Consolandich Cirisola, María Victoria García Olazabal, Laura Hermida Bruno, Federico Todeschini Safi, Lucia Piriz Trindade, Priscila Larcher Longo, Maria Cristina Chavantes, Cinthya Cosme Gutierrez Duran, Kristianne Porta Santos Fernandes, Raquel Agnelli Mesquita Ferrari, Sandra Kalil Bussadori, Lara Jansiski Motta, Anna Carolina Ratto Tempestini Horliana). Universidad Católica del Uruguay (UCU), Montevideo, Uruguay. Universidade São Judas Tadeu (USJT), São Paulo, Brazil. Universidade Brasil, São Paulo, Brazil (Ricardo Scarparo Navarro). These authors contributed equally to this work (María Victoria García Olazabal, Laura Hermida Bruno, Federico Todeschini Safi, Lucia Piriz Trindade, Priscila Larcher Longo, Ricardo Scarparo Navarro, Anna Carolina Ratto Tempestini Horliana). These authors also contributed equally to this work (Luis Eduardo Pascuali Moya, Rolf Wilhem Consolandich Cirisola, Maria Cristina Chavantes, Cinthya Cosme Gutierrez Duran, Kristianne Porta Santos Fernandes, Raquel Agnelli Mesquita Ferrari, Sandra Kalil Bussadori, Lara Jansiski Motta). **data availability Statement-** all data will be available for the readers. **funding Statement –** The project receives a grant from the National Council for Scientific and Technological Development process (CNPq:3146682020-9). **ethics approval Statement -** The project received approval from the Research Ethics Committee of Universidad Católica del Uruguay (UCU), process: 221014b. **patient consent Statement –** All the participants included signed an Informed consent. **clinical trial registration:** NTC05924204 (version 08/2023).

## Abstract

Recent studies have shown that photobiomodulation (FBM) can modulate pain after the placement of elastomeric separators, however, to date, there is no ideal protocol for its application. Therefore, the objective of this study will be to evaluate the effect of photobiomodulation on pain control 24 hours after the placement of elastomeric separators using the visual analog scale (VAS). Twenty-five participants between 13 and 30 years old with the need for the placement of orthodontic bands in the lower first molars bilaterally will be included, which establishes a sample of fifty molars (right and left). Elastomeric separators will be placed on the mesial and distal surfaces of the right and left molars. The study groups will be G1 (experimental) - elastomeric separators + FBM (diode laser, 808nm, 100mw power, with 4 J, 3 points per vestibular and 3 points per palatal, single session) and G2-(control)-elastomeric separators + FBM simulation. Treatment will be randomized to the right molar and the opposite treatment will be applied to the left side. The patient and the evaluator will be blinded to the intervention performed. The primary outcome variable will be spontaneous pain assessed 24 hours after the placement of elastomeric separators measured with the VAS scale. Secondary outcome variables will be pain during mastication (measured with the VAS scale) at 72h after the placement, count of the number of analgesics (paracetamol), and local temperature (measured with a digital thermometer). To assess the impact of oral health on quality of life. of the participant, the OHIP-14 questionnaire will be applied. All the outcomes will be evaluated at baseline, 24 and 72 hours after the placement of elastomeric separators. If the data are normal, they will be submitted to the ANOVA – one-way test. Data will be presented as means ± SD and the p-value will be set to < 0.05.

## Introduction

The demand for orthodontic treatment has increased exponentially in recent decades (Pacheco-Pereira et al., 2015), however, pain remains a limiting factor for this treatment (Eslamian et al., 2014). The placement of elastomeric separators is strongly associated with pain and discomfort (Borzabadi-Farahani et al., 2017). Patients report that the placement of elastomeric separators is the most painful part of the entire treatment (Farias et al., 2017). It is reported that 90% of patients undergoing orthodontic treatment experience varying degrees of pain after placement of elastomeric separators, initial wire insertions, and activations. (Qamrudin et al., 2014). In general, it starts after 4 hours of inserting the separator, reaches its peak within 24 hours, remains uncomfortable for the next 3 days, and subsides and dissipates in approximately 6 to 8 days (Abtahi et al., 2013)

The tooth movement generated is a biological process caused by forces that are transmitted through biochemical signals and its answer depends on the physiology of the tissues (Krishnan et al., 2009). Cells and interleukins participate in this process through various interactions (Jeon et al., 2021) causing acute inflammation in the periodontal tissue (Jeon et al., 2021; Li et al., 2018). As the pain is generated from an inflammatory process, non-steroidal anti-inflammatory drugs (NSAIDs) are the gold standard to reduce discomfort (Kohli et al., 2011). However, in addition to the systemic adverse effects of these medications, caution must be considered of their influence on decelerating rate movement of the teeth. (Qamruddin et al., 2014)

Photobiomodulation is effective in controlling pain in several areas of dentistry, however, until now there is no ideal application protocol for its reduction after the placement of elastomeric separators. A systematic review showed that the laser application method and the laser parameters were widely varied in tested protocols (Farzan et al., 2014). Establishing ideal parameters for pain reduction with laser is the objective of several authors (Qamruddin et al., 2014, Abtahi et al., 2013, Borzabadi-Farahani et al., 2017) and meanwhile, more evidence is necessary to define the ideal dosimetric protocol to mitigate pain after placing elastomeric separators (Farzan et al., 2014). The most used protocol includes diode, such as aluminum arsenic, and gallium applied continuously and in direct contact with irradiated areas in the near-infrared range (Farzan et al., 2014). It was also observed that double irradiation did not present an advantage over irradiation in a single dose, also, that is observed that it is better to start the treatment as soon as possible, preferably immediately after placing the elastomeric separators (Farzan et al., 2014). However, more evidence is needed to determine the best dosimetry and the best intervention protocol. Therefore, the primary objective of the study is to assess whether photobiomodulation can promote pain control (spontaneous and during mastication) immediately after placing elastomeric separators through the visual analog scale (VAS).

## Methods

This randomized, controlled, double-blind, split-mouth clinical trial meets the criteria for designing a clinical study as outlined in the SPIRIT Statement (Figure 1). The study received approval from the Committee for Ethics in Research (CEP) at the Universidad Católica del Uruguay (#221014b) on 14^th^ October 2022. Participants will be recruited from patients undergoing orthodontic treatment at the Orthodontic Clinic of the Specialization Course in Orthopedics and Orthodontics at the Catholic University of Uruguay, located in Montevideo, Uruguay. This treatment involves the installation of bands on the lower first permanent molars (both left and right). We have not prospectively recruited human participants for the study (conducted a clinical trial, distributed questionnaires, or obtained tissues, data, or samples for the purposes of this study). The study will start in 30^th^ September and will finish in 30th July 2024. This study does not include minors. All the participants provided informed consent (written and verbal). This document was approved by our Ethics Committee. This study are not reporting a retrospective study of medical records or archived samples.

**Figure 1.**
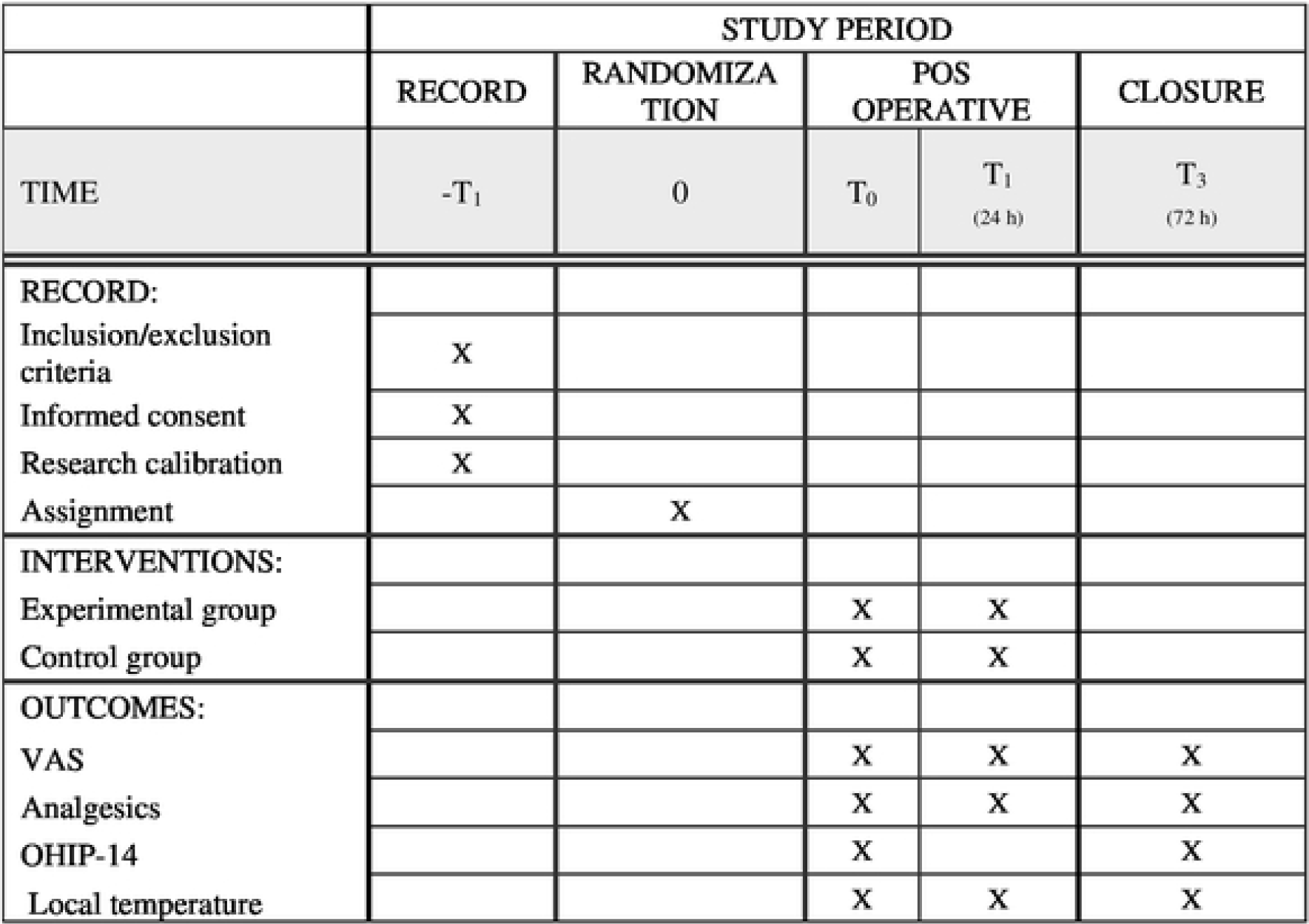

Any future changes to the study will be promptly reported to the CEP and will be disclosed in subsequent publications. A signed copy of the informed consent form will be provided to each participant via email. The principal researcher will apply the IC. The project has been registered on the ClinicalTrials.gov website (NTC05924204). No conflicts of interest exist regarding any products used in the study or any authors involved. The study data will be published without any restrictions on data inclusion. All data will be available for consultation, and participants will have access to their medical records at any time. All data will be managed by the principal researcher on a computer with password and no internet.

### Description of the sample

This investigation will be realized in the Orthodontic Department of the Clinic associated with the Specialization Course in Orthopedics and Orthodontics at the Catholic University of Uruguay. The study will include individuals requiring metallic devices by the Roth prescription, along with the necessity for the placement of an orthodontic band.

### Inclusion/exclusion criteria

Participants’ inclusion criteria are:

- Need to place bilateral orthodontic bands on the lower first molars.
- Age 13 to 30 years old.
- Both genders
- No comorbidities.
- Never used orthodontic appliances.
- Preserved surrounding spaces.
- Healthy permanent dentition with good hygiene.

Participants’ exclusion criteria are:

- Who are taking drugs that affect bone metabolism and the inflammatory process (for example corticosteroids, bisphosphonates), who have used anti-inflammatory drugs or analgesics for less than 1 week since placing elastomeric separators.
- Present general pathologies (craniofacial anomalies) and local ones (dental anomalies) that affect the result of the movement of the teeth,
- Smokers, embarrassed, or breastfeeding women.
- Allergic to paracetamol
- Use of any drugs, different from those prescribed by investigators

### Sample size calculation

The sample size to evaluate the analgesic effect of PBM was determined based on previous studies by Qamruddin et al., 2016 using the G*Power software (version 3.1.9.2). An average spontaneous pain of 5 was considered on the VAS scale in the control group and 2 in the laser treatment group after 24 hours. The highest standard deviation found (±3.24) was used. Considering a significance level of 5%, test power of 80%, and size effect of 0.60 for the detection of differences between groups, a total of 25 patients will be required for each group which totalizes 50 patients.

### Training of examiners

An examiner, with experience in dentistry and specialization in orthodontics, will practice the visual analog scale.

### Randomization

For randomization, a program available on the internet and a random sequence generator (https://www.sealedenvelope.com/) will be used. The treatment for the molar on the right side will be randomized, and on the left side, the opposite intervention will be performed (Figure 2). Opaque envelopes will be identified with sequenced numbers within the corresponding group (experimental or control) and will be introduced according to the generated random sequence. The envelopes will be sealed and remain sealed in a safe place until the end of the statistical analysis. The generation of the random sequence and the preparation of the envelope will be carried out by a person not directly involved in the study.

**Figure 2.**
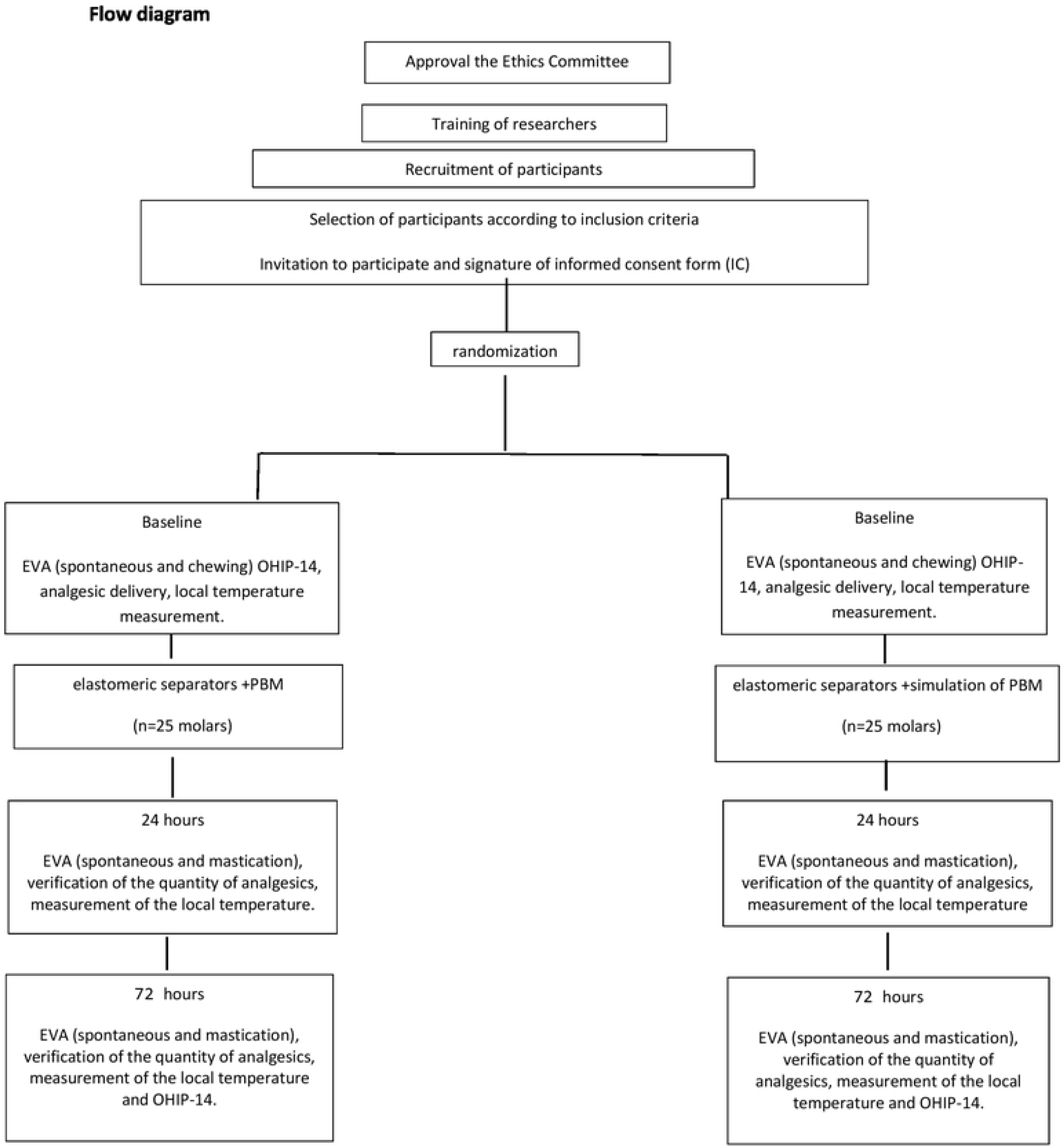

### Evaluations before treatment

Firstly, the participants who are interested in participating in the study will read the informed consent (IC) and the investigator will perform a verbal explanation. If they accept to participate, they will sign the term (IC) and fill anamnesis form. The researcher will apply the OHIP-14 questionnaire.

### Placement of elastomeric separators

Each elastomeric separator (AO Sheboygan, Wisconsin), will be placed with a dental hilum (Oral B; Procter & Gamble, Cincinnati, Ohio) between the mesial and distal interproximal contacts of the first permanent lower molars on both sides, always starting from the right side.

Before any intervention, the investigator responsible for applying the FBM will remove and open 1 envelope (without changing the numerical sequence of envelopes). Thus the investigator will undergo the intervention according to the assigned group, starting always on the right side. On the left side, the treatment will be the opposite of the one designated by randomization.

Thus, the 25 participants (50 bilateral molars) will be randomly assigned to groups G1 or G2:

#### Treatment group

Photobiomodulation (n=25 lower molars): Immediately after placing the elastomeric separators, the photobiomodulation will be performed as follows: 3 applications per buccal and 3 applications perlingual on the first lower molars, in a single session. Two applications will be made on the cervical third (on the mesial and distal side of the molar primer) and one application on the apical third of the roots. (Figure 3). A Ga-Al-As laser (Therapy XT, DMC Equipamentos, São Carlos, Brazil) will be used with an output power of 100 mW. The tip of the device will be placed perpendicular to the mucosa, in contact, but without exerting pressure. Each point will be irradiated with 4 J for 40 seconds, totaling 24 J per tooth (12 J on the buccal side and 12 J on the lingual side). Table 1 shows the dosimetric parameters, based on a previous study (Ortega, 2019).

**Table 1:** Dosimetric parameters (Ortega et al, 2019)

| Parameters | Values/Treatment |
| --- | --- |
| Wavelength [nm] | 808 |
| operating mode | Continuous |
| Radiant power [mW] | 100 |
| Irradiance [mW/cm<sup>2</sup>] | 35 |
| Surface area [cm<sup>2</sup>] | 0,002826 |
| Exposure time [s] | 40 (each point) |
| Radiant exposure [J/cm<sup>2</sup>] | 1.415 |
| Radiant energy [J] | 4 |
| Number of irradiated points | 6 points per tooth: |
|  | Vestibular (2 cervical and 1 apical) |
|  | Palatine (2 cervical and 1 apical) |
| application technique | In contact, at 90 degrees with the surface |
| Number of sessions and frequency | Single session, immediately after placing the elastomeric separator |

**Figure 4.**
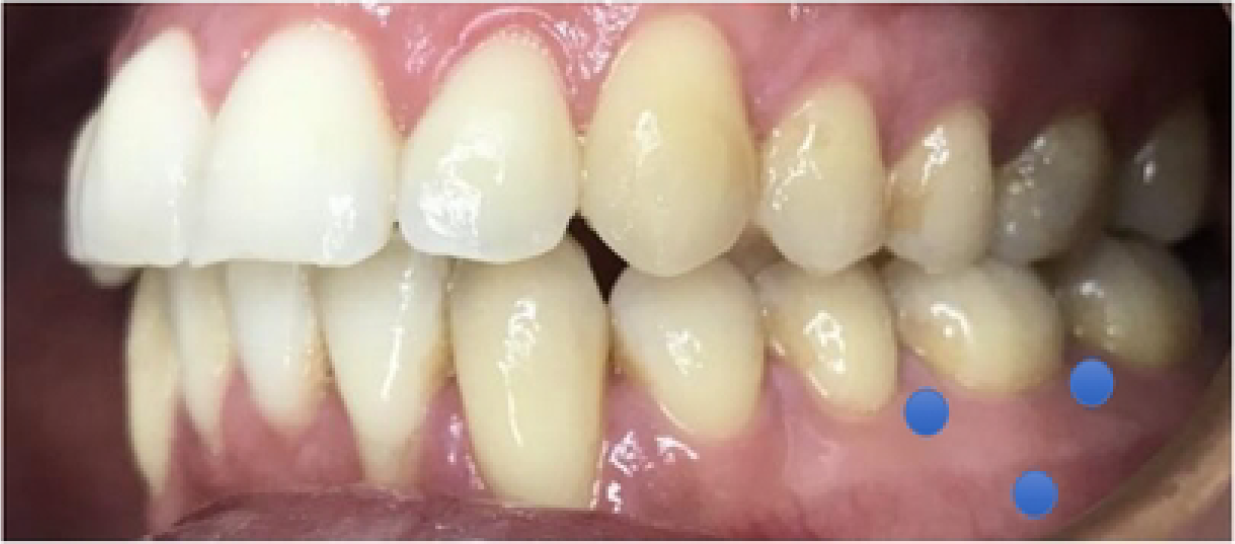
Laser application regions on the vestibular face. In the same position, lingual irradiation will be carried out (source: case of the Clinica de Salud UCUDAL)

The description of the equipment, the dosimetric parameters, and the number of FBM applications are described in Table 1

#### Control Group

Simulation of Photobiomodulation (n=25 lower molars): The teeth that will receive the FBM simulation will be treated identically to the G1 group. The person responsible for applying FBM will simulate the irradiations by placing the devices in the same places described for the FBM group, however, the equipment will remain off. The sound of the devices will be simulated to blind the intervention.

There are no criteria for discontinuing, because no adverse effects are expected

### Blinding

The research team will be:

- Orthodontist: who supervises all the procedures and does not know in which teeth the FBM is being applied.
- Examiner - (will carry out all the analysis of the results, apply the visual analog scale and evaluate the number of drugs ingested. This investigator will be aware of the interventions carried out in the groups).
- Investigator who will perform the intervention (or simulation) of FBM in patients. Only this investigator will know all the nature of interventions. The identification of each group will be revealed by this researcher after the statistical analysis of the data.
- Estatistic– will be blind regarding the treated groups (she will receive the sheets with G1 and G2 information only).
- Participants - will not know the interventions performed because the FBM will be simulated in the control group.

### Outcomes

- Pain - through the Visual Analog Scale (VAS) at the beginning, 24 hours after FBM (primary outcome), and 72 hours after FBM (secondary outcome). Patients will report their perception of pain by marking their level of discomfort on the visual analog scale (VAS), which ranges from 0 (absence of pain) to 10 (unbearable pain).
- Number of analgesics taken within a period of 3 days. The analgesic used will only be paracetamol. Participants will be instructed to use the medication if necessary and they will be given a paracetamol blister.
- The quality of life-related to oral health will be evaluated using the OHIP-14 questionnaire at the beginning and 3 days after treatment.
- Local temperature - evaluated using a digital thermometer in baseline, 24 h, and 72 h after treatment.

### Statistical analysis

Initial descriptive analyses will be performed, considering all the variables measured in the study, both quantitative (mean and standard deviation) and qualitative (frequencies and percentages). Subsequently, normality analyses will be conducted. Based on the results of these tests, appropriate statistical tests will be selected for each specific analysis. A significance level of 5% probability or the corresponding p-value will be used for all tests. All analyses will be executed using the statistical software SPSS for Windows, version 9.1.

## Data Availability

Deidentified research data will be made publicly available when the study is completed and published.

## Notes

**conflict of interest disclosure** All authors disclose any financial and personal relationships with other people or organizations that could inappropriately influence (bias) this study.

### Competing Interest Statement

The authors have declared no competing interest.

### Clinical Trial

NCT05924204

### Clinical Protocols

https://clinicaltrials.gov/study/NCT05924204?term=NCT05924204&rank=1

### Funding Statement

The funders did not and will not have a role in study design, data collection and analysis, decision to publish, or preparation of the manuscript.

### Author Declarations

The study received approval from the Committee for Ethics in Research (CEP) at the Universidad Cat#x00F3;lica del Uruguay (#221014b) on 14th October 2022.

## REFERENCES

Pachêco-Pereira C, Pereira JR, Dick BD, Perez A, Flores-Mir C. Fatores associados à satisfação do paciente e dos pais após tratamento ortodôntico: revisão sistemática. Am J Orthod Orntofacial Orthop. 2015 Out;148(4):652–9. doi: 10.1016/j.ajodo.2015.04.039.26432321.

Eslamian L, Borzabadi-Farahani A, Hassanzadeh-Azhiri A, et al. The effect of 810-nm low-level laser therapy on pain caused by orthodontic elastomeric separators. Lasers Med Sci 2014;29:559–64.

Borzabadi-Farahani A, Cronshaw M. Lasers in orthodontics. Lasers in Dentistry—Current Concepts. Springer; 2017. p. 247–71. doi:10.1007/978-3-319-51944-9_12

Farias RD, Motta RH. Low-level laser therapy for controlling pain in orthodontic patients during the use of elastic separators: a randomized clinical trial. Laser Phys Lett. 2018;15(9):095602. doi:10.1088/1612-202x/aad1c1

Qamruddin I, Alam MK, Fida M, Khan AG. Effect of a single dose of low-level laser therapy on spontaneous and chewing pain caused by elastomeric separators. Am J Orthod Dentofacial Orthop. 2016 Jan;149(1):62–6. doi: 10.1016/j.ajodo.2015.06.024. PMID: 26718379.

Abtahi SM, Mousavi SA, Shafaee H, Tanbakuchi B. Effect of low-level laser therapy on dental pain induced by separator force in orthodontic treatment. Dent Res J (Isfahan). 2013 Sep;10(5):647–51. PMID: 24348624; PMCID: PMC3858741

Krishnan V, Davidovitch Z. Em um caminho para a implantação dos mecanismos biológicos do movimento ortodôntico do dente. J Dent Res. 2009; 88:597–608. doi: 10.1177/00222034509338914. [PubMed] [CrossRef] [GoogleScholar]

Jeon HH, Teixeira H, Tsai A. Visão mecanicista do movimento odontológico ortodôntico com base em estudos em animais: Revisão crítica. J Clin Med. 2021 Abr 16;10(8):1733. doi: 10.3390/jcm10081733.33923725; PMCID: PMC8072633.

Li Y, Jacox LA, Little SH, Ko CC. Movimento Odontológico Ortodôntico: Biologia e Implicações Clínicas. Kaohsiung J Med Sci. 2018 Abr;34(4):207–214. doi: 10.1016/j.kjms.2018.01.007. Epub 2018 Feb 3. 29655409.

Kohli SS, Kohli VS. Effectiveness of piroxicam and ibuprofen premedication on orthodontic patients’ pain experiences: a randomized control trial. Angle Orthod 2011;81:1097–102.

Farzan A, Khaleghi K. The Effectiveness of Low-Level Laser Therapy in Pain Induced by Orthodontic Separator Placement: A Systematic Review. J Lasers Med Sci. 2021 Jun 24;12:e29. doi: 10.34172/jlms.2021.29. PMID: 34733752; PMCID: PMC8558704.

Ortega SM, Gonçalves MLL, da Silva T, Horliana ACRT, Motta LJ, Altavista OM, Olivan SR, Santos AECGD, Martimbianco ALC, Mesquita-Ferrari RA, Fernandes KPS, Bussadori SK. Evaluation of the use of photobiomodulation following the placement of elastomeric separators: Protocol for a randomized controlled clinical trial. Medicine (Baltimore). 2019 Oct;98(43):e17325. doi 10.1097/MD.0000000000017325. PMID: 31651838; PMCID: PMC6824799.

